# The inevitability of Covid-19 related distress among healthcare workers: findings from a low caseload country under lockdown

**DOI:** 10.1101/2020.06.14.20130724

**Authors:** Feras Ibrahim Hawari, Nour Ali Obeidat, Yasmeen Izzat Dodin, Asma Salameh Albtoosh, Rasha Mohammad Manasrah, Ibrahim Omar Alaqeel, Asem Hamza Mansour

## Abstract

**Objectives:** To characterize psychological distress and factors associated with distress in healthcare practitioners working during a stringent lockdown in a country (Jordan) with one of the lowest incidence rates of Covid-19 globally.

**Methods:** A cross-sectional online survey sent to physicians, nurses and technicians, and pharmacists working in various hospitals and community pharmacies. Demographic, professional and psychological characteristics (distress using Kessler-6 questionnaire, anxiety, depression, burnout, sleep issues, exhaustion) were measured as were potential sources of fear. Descriptive and multivariable statistics were performed using level of distress as the key outcome.

**Results:** We surveyed 1,006 practitioners (55.3% females). Approximately 63%, 13%, 17% and 7% were nurses/technicians, physicians, pharmacists, and other nonmedical personnel (respectively). 32% suffered from high distress while 20% suffered from severe distress. Exhaustion, anxiety, depression, and sleep disturbances were reported (in past seven days) by approximately 34%, 34%, 19%, and 29% of subjects (respectively). Being older or male, perception of effective protective institutional measures, and being satisfied at work, were significantly associated with lower distress. Conversely, suffering burnout; reporting sleep-related functional problems; exhaustion; being a pharmacist (relative to a physician) and working in a cancer center; harboring fear about virus spreading; fear that the virus threatened life; fear of alienation from family/friends; and fear of workload increases, were significantly associated with higher distress.

**Conclusion:** Despite low caseloads, Jordanian practitioners still experienced high levels of distress. Identified demographic, professional and psychological factors influencing distress should inform interventions to improve medical professionals’ resilience and distress likelihood, regardless of the variable Covid-19 situation.

## INTRODUCTION

Healthcare practitioners (physicians, nurses, medical technicians, and pharmacists) globally are currently facing extraordinary circumstances as a result of the Covid-19 pandemic. From the world’s past experiences with other viral outbreaks such as SARS, it is evident that such circumstances impact healthcare practitioners’ mental as well as physical well-being, with carry-over effects also being reported even after resolution of outbreaks.(1-3) For example, practitioners in past outbreaks have reported depression, anxiety, burnout and fatigue during and following these difficult times.(4-6) The experience with Covid-19 is no different, if not more pronounced, due to its being more widespread: the on-going outbreak will have dramatic short as well as long-term mental health consequences across various subgroups of the population (patients, providers and the lay public).(7)

Globally, it has been found that healthcare practitioners in countries such as China, Iran, the US and Italy have suffered from heightened anxiety, depression, stress and insomnia during the Covid-19 outbreak.(8-12) These countries share the fact that the Covid-19 outbreak exerted a considerable toll, given that the caseload in these countries was high. Converse to this, the Kingdom of Jordan in the Middle East represents a differing situation and an interesting case study: the country has recorded some of the lowest numbers of cases globally due to its early and firm response to the outbreak. Stringent measures were put in place promptly in March, including border closures and limiting free travel; testing and enforced 14-day isolation of all in-bound travelers in designated hotels (or hospitals if testing positive for Covid-19), followed by an additional 14-day quarantine after leaving those hotels or being discharged from hospitals; imposing a six-week lockdown proceeded by a staggered re-opening of select sectors; banning social gatherings; and restricting the public’s movement using a daily curfew.(13) As of June 7, 2020, the country of approximately ten million inhabitants had recorded 795 cases and 9 deaths.(14) Like other countries, frontline workers including healthcare practitioners, have been a key component of the country’s response plan.

Despite their key roles in controlling the outbreak, little has been published about Jordanian frontline workers’ experiences and mental health. Specifically in the context of Jordanian healthcare workers, some studies examined knowledge and readiness as it pertains to Covid-19, in pharmacists, dentists and physicians.(15-17) One study examined general anxiety and depression of a national sample inclusive of healthcare practitioners.(18) None have examined in an in-depth manner the prevalence and sources of distress in this group, within its unique local context. Evaluating the predisposition of practitioners to distress, anxiety, sleep and burnout is critical in order to identify mechanisms to address and hopefully alleviate such stress.(19, 20) Importantly, understanding how distress can vary across different scenarios of Covid-19 spread, regardless of caseload, provides valuable information about how healthcare practitioners will potentially respond to the continually changing Covid-19 circumstances across the world.

We sought to evaluate Jordanian healthcare practitioners (physicians, nurses and technicians, pharmacists) fear, distress, anxiety, depression, sleep quality, and fatigue during the period when the country was on high-alert, implementing stringent national measures to control the outbreak. Published studies on healthcare worker distress have been generated from countries with a high caseload. We hypothesized that despite a low caseload, distress, fear and anxiety would nevertheless be prevalent among healthcare workers as a result of the potential threat of disease emergence or spread. We also hypothesized that key factors, namely, demographics such as age and gender, profession (particularly professions that experienced greater service demand during the outbreak), and workplace environment would be significantly associated with distress. In addition, we measured reported availability of personal protective equipment (PPE) during the country’s Covid-19 lockdown. Our study thus aimed to shed light on a low caseload setting and provide a unique perspective on healthcare worker reactions and understand which factors could predispose them to a heightened sense of distress.

## METHODS

### Study design and sample

A cross-sectional Arabic online survey (https://www.questionpro.com/) was developed and distributed across key governmental and academic hospitals and in community pharmacies largely in the Central region of the country (during lockdown, only hospitals and community pharmacies continued their operations). Distribution channels were purposeful, targeting physicians, nurses, technicians, and pharmacists. Channels included email, text-messaging, and social media groups restricted to healthcare professionals potentially working in these key institutions. The questionnaire was available between April 21, 2020 and May 17, 2020.

### Study variables and measures

The questionnaire was developed and reviewed by a core team of medical staff involved in both research and Covid-19 screening and potential management; and was approved by an AAHRPP (Association for the Accreditation of Human Research Protection Programs, Inc) accredited Institutional Review Board. It was composed of the following sections:

i. Mental and general physical health: distress (using the Kessler 6 scale);(21) burnout (using a single-item measure);(22) anxiety (Patient-Reported Outcomes Measurement Information System – PROMIS – Anxiety short-form);(23) depression (PROMIS depression short-form);(24) sleep (using three items from the PROMIS sleep-related impairment and the PROMIS sleep impact short forms);(25, 26) and fatigue (using two items from the PROMIS Fatigue short-form).(27) Our primary outcome of interest was the Kessler distress score [in the past 30 days], which was divided into four categories of no distress (score of 0), low distress (scores of 1 to 5), moderate distress (scores of 6 to 10), and high distress (scores of 11 to 24).(28) The Kessler 6 scale was selected due to its brevity and reliability, and due to its appropriate reference time period of 30 days, which would have captured most of the lockdown period. Rough cut-offs of 11 (from a total score of 20) were used to identify at least moderate anxiety or depression (this cut-off was roughly selected as it equates to the T-score that has been shown to be approximately equivalent to other anxiety and depression measure cut-offs (29, 30)).
ii. Fear – various items covering potential sources of distress (fear) due to the Covid-19 outbreak were adapted from other studies that were conducted in comparable situations, namely the SARS outbreak.(1, 2) Fear statements were originally measured using a 5-point Likert scale (from “not at all” to “a very great extent”) and then dichotomized for analysis, by considering those who responded “to a considerable extent” and “to a very great extent” as fearful regarding the statement.
iii. Workplace characteristics and perceptions about working environment (a selection of items were adapted from other studies).(31)
iv. Limited access to personal protective equipment (PPE) in the workplace was investigated in our study as a potential source of distress given the global shortage of care resources, including PPE, amid the Covid-19 pandemic.(32) Availability of specific personal protective equipment and perceived protection imparted by these equipment were measured (items were adapted from a previous SARS-related study).(33) We explored individual equipment and also created a summary variable, ‘PPE availability’, which was defined as having access to a mask (surgical or N95), gloves, a gown, and shoe covers.
v. Demographic and professional characteristics.

### Statistical analysis

Descriptive bivariate statistics were first conducted to characterize levels of distress, fear, anxiety and depression. We specifically focused, in our primary analysis, on examining whether or not distress varied across demographic and professional characteristics, its association with other measures of mental health (such as burnout, fatigue, anxiety and depression), and the potential sources of fear associated with overall distress. To further understand the ways in which the various mental health related, demographic and professional characteristics were associated with distress, multivariable ordinal logistic regression analysis was conducted to identify significant factors that were associated with an increased odds of being in a higher distress category.

Covariate selection was generally based on clinical judgement as well as statistical findings during bivariate analysis to examine which factors correlated with distress levels. With regards to fear-related and PPE-related items, because they were numerous and to maintain model parsimony, we first simultaneously evaluated fear items and distress, and PPE-related items (availability and perceived effectiveness of each PPE item) and distress; and included only significant fear items and PPE-related items in the model (therefore, only items related to surgical masks and N95 masks were retained).

In secondary analyses, we examined the availability of PPE as well as the perceptions of respondents with regards to their protective effect. We specifically examined these in the context of healthcare profession, given that community pharmacists operate in a different working environment than pharmacists, nurses and physicians at hospitals.

All analyses were conducted in STATA 16.(34, 35)

## RESULTS

### Primary analysis

Our final sample included 1,006 Jordanian healthcare practitioners with a mean age of 33.5 years (range 21 to 67), and was comprised of 55.3% females. With regards to profession, 63.5% of the respondents were nurses and technicians, 12.7% were physicians, and 16.8% were pharmacists (7.0% fell in “other” categories of nonmedical personnel in the health sector). Roughly 40% of respondents worked in a government or academic hospital that provided diagnostic (but not treatment) services for Covid-19; 4.0% worked in a government or academic hospital that provided treatment services for Covid-19; 45.0% worked in a specialized cancer center (which was also authorized to conduct Covid-19 diagnosis); and 11.5% worked in community pharmacies. About 20% of the sample suffered from very severe distress (13 or higher Kessler-6 score). When Kessler scores were categorized into four levels, 31.5% reported high levels of distress. In the past seven days [from survey completion], approximately 34% and 19% also reported at least moderate anxiety and depression (respectively). About 29% reported sleep issues (trouble falling asleep or staying up atleast half the night), 54% of whom also experienced problems functioning during the day as a result; and 34% reported considerable exhaustion, 56% of whom also experienced problems functioning during the day as a result. Descriptive statistics of the sample, in relation to reported levels of distress, are presented in Table 1. Females and younger respondents were more likely to fall in the higher distress levels (than males and older respondents); respondents in higher distress level categories were more likely to live with older people, whereas respondents falling in lower distress levels were more likely to be married and have children. Professional characteristics associated with higher distress included having fewer years of experience, working with suspected Covid-19 cases, and experiencing a high workload in the past 30 days. Having either less or more than a Bachelors degree were both associated with lower distress, as were reported PPE availability, being satisfied in the general workplace, and reporting effective institutional safety measures in place. Suffering burnout, exhaustion or sleep problems all were significantly associated with higher distress.

**Table 1.** Demographic, professional and workplace characteristics across distress levels in a sample of Jordanian healthcare practitioners (n=1,006). Column total percentages presented.

|  | No distress<br>(n=35) | Low distress<br>(n=311) | Moderate<br>distress<br>(n=343) | High distress<br>(n=317) | P-value |
| --- | --- | --- | --- | --- | --- |
| **Age (mean)** | 41.6 | 35.9 | 33.0 | 30.9 | 0.000 |
| **Gender (being male)** | 23 (65.7%) | 170 (54.7 %) | 148 (43.2 %) | 109 (34.4 %) | 0.000 |
| **Live with spouse (yes, relative to no)** | 28 (82.4%) | 219 (71.1%) | 225 (65.8%) | 171 (54.29%) | 0.000 |
| **Have children (yes, relative to no)** | 26 (74.3 %) | 195 (62.7%) | 197 (57.4%) | 156 (49.2%) | 0.001 |
| **Live with old people (yes, relative to no)** | 15 (42.9%) | 128 (41.2 %) | 139 (40.5%) | 166 (52.4 %) | 0.009 |
| **Live with young people (yes, relative to no)** | 29 (82.9%) | 241 (77.5%) | 271 (79.0%) | 258 (81.4%) | 0.626 |
| **Education level** |  |  |  |  | 0.002 |
| Diploma or less | 9 (25.7%) | 50 (16.1%) | 44 (12.8%) | 31 (9.8%) |  |
| Bachelor degree | 16 (45.7%) | 199 (64.0%) | 253 (73.8%) | 231 (72.9%) |  |
| Masters, PhD | 10 (28.6%) | 62 (19.9%) | 46 (13.4%) | 55 (17.4%) |  |
| **Occupation (n=990)\*** |  |  |  |  | 0.023 |
| Nurses and technicians | 22 (64.7%) | 196 (64.9%) | 209 (61.8%) | 202 (63.9%) |  |
| Physicians | 6 (17.7%) | 42 (13.9%) | 42 (12.4%) | 36 (11.4%) |  |
| Pharmacists | 0 (0.0%) | 40 (13.3%) | 65 (19.2%) | 61 (19.3%) |  |
| Other (Non-medical staff) | 6 (17.7%) | 24 (8.0%) | 22 (6.5%) | 17 (5.4%) |  |
| **Years of experience in the field (mean)** | 17.2 | 12.0 | 9.5 | 8.1 | 0.000 |
| **Site of work** |  |  |  |  | 0.251 |
| ICU & ER | 9 (25.7%) | 82 (26.6%) | 91 (26.7%) | 86 (27.4%) |  |
| Hospital medical departments | 24 (68.6%) | 181 (58.8%) | 194 (56.9%) | 189 (60.2%) |  |
| Community pharmacies | 0 (0.0%) | 35 (11.4%) | 46 (13.5%) | 36 (11.5%) |  |
| Other (Hospital non-medical departments) | 2 (5.7%) | 10 (3.3%) | 10 (2.9%) | 3 (1.0%) |  |
| **Type of institution (government or academic)** |  |  |  |  | 0.183 |
| Specialized hospital (cancer) | 19 (54.3%) | 130 (42.1%) | 158 (46.1%) | 145 (46.2%) |  |
| Non-cancer/general hospital | 16 (45.7%) | 145 (46.9%) | 138 (40.2%) | 135 (43.0%) |  |
| Community pharmacy | 0 (0.0%) | 34 (11.0%) | 47 (13.7%) | 34 (10.8%) |  |
| **Exposed to potential COVID patients in line of work (yes, relative to no)** | 12 (34.3%) | 129 (41.5%) | 159 (46.4%) | 170 (53.6%) | 0.008 |
| **Work in a Covid-19 specialized ward** | 3 (8.6%) | 54 (17.4%) | 45 (13.1%) | 52 (16.4%) | 0.284 |
| **Experienced a high workload during past 30 days (yes, relative to no)** | 6 (17.1%) | 67 (21.5%) | 111 (32.4%) | 141 (44.5%) | 0.000 |

|  | **No distress**<br>**(n=35)** | **Low distress**<br>**(n=311)** | **Moderate**<br>**distress**<br>**(n=343)** | **High distress**<br>**(n=317)** | **P-**<br>**value** |
| --- | --- | --- | --- | --- | --- |
| **Was satisfied at work (agree,**<br>**relative to all other responses)** | 33 (94.3%) | 284 (91.6%) | 254 (74.1%) | 160 (50.6%) | 0.000 |
| **Agreed that place of work**<br>**implemented effective safety**<br>**measures** | 30 (90.9%) | 214 (73.3%) | 202 (62.0%) | 144 (47.7%) | 0.000 |
| **Reported surgical masks were**<br>**available** | 28 (84.9%) | 234 (80.1%) | 237 (72.7%) | 201 (66.6%) | 0.001 |
| **Reported N95 masks were**<br>**available** | 22 (66.7%) | 142 (48.6%) | 144 (44.2%) | 93 (30.8%) | 0.000 |
| **Reported eye guards were**<br>**available** | 20 (60.6%) | 126 (43.2%) | 131 (40.2%) | 91 (30.1%) | 0.000 |
| **Reported gowns were available** | 28 (84.9%) | 218 (74.7%) | 201 (61.7%) | 182 (60.3%) | 0.000 |
| **Reported gloves masks were**<br>**available** | 32 (97.1%) | 263 (90.1%) | 287 (88.0%) | 246 (81.5%) | 0.003 |
| **Reported shoe covers were**<br>**available** | 26 (78.8%) | 212 (72.6%) | 201 (61.7%) | 162 (53.6%) | 0.000 |

Specific sources of fear and concerns that were prevalent in our sample included (Table 2): fear of respondents infecting others (the overwhelming majority, 83.2%, reported this), and fear of families becoming infected in general (64.5% were concerned). Conversely, only 32.7% were concerned about themselves being infected. Other sources of fear that resonated with the sample included financial concerns as a result of the outbreak (56.6%); concerns about other health problems in the family as a result of the outbreak (51.5%); fear about their own susceptibility to the virus (virus is nearing, 43.2%). Approximately 35% were concerned about increasing workloads or being quarantined as a result of the outbreak. Table 2 displays the various fear items that were of most relevance to respondents by distress level, as well as raw scores for anxiety and depression in the sample. Distress levels correlated consistently and significantly with all fear items and with anxiety and depression scores (Table 2).

**Table 2.** State of fears and mental health across distress levels in a sample of Jordanian healthcare practitioners. Column total percentages presented.

|  | **No distress**<br>**(n=35)** | **Low**<br>**distress**<br>**(n=311)** | **Moderate**<br>**distress**<br>**(n=343)** | **High**<br>**distress**<br>**(n=317)** | **P-**<br>**value** |
| --- | --- | --- | --- | --- | --- |
| **Anxiety, past 7 days raw score (mean)** | 4.2 | 6.3 | 9.1 | 12.5 | 0.000 |
| **Depression, past 7 days raw score (mean)** | 4.1 | 4.7 | 6.3 | 11.1 | 0.000 |
| **Experienced [quite a bit, very much] sleep disturbances (reference: those who reported some or none)** | 1 (2.9%) | 39 (12.5%) | 78 (22.7%) | 172 (54.3%) | 0.000 |
| **Had [quite a bit, very much] exhaustion (relative to those who reported some or none)** | 1 (2.9%) | 36 (11.6%) | 98 (28.6%) | 204 (64.4%) | 0.000 |
| **Had atleast one symptom of burnout (relative to those with no symptoms)** | 2 (5.7%) | 30 (9.7%) | 94 (27.4%) | 204 (64.4%) | 0.000 |
| ***Fear items*** |  |  |  |  |  |
| High level of fear of being infected | 4 (11.4%) | 54 (17.4) | 115 (33.5%) | 156 (49.2%) | 0.000 |
| High level of fear of infecting others | 20 (57.1%) | 226 (72.7%) | 297 (86.6%) | 294 (92.7%) | 0.000 |
| Felt virus was close and they were susceptible | 6 (17.1%) | 88 (28.3%) | 147 (42.9%) | 194 (61.2%) | 0.000 |
| Felt life was under threat | 0 (0.0%) | 40 (12.9%) | 90 (26.2%) | 159 (50.2%) | 0.000 |
| Felt virus was going to go out of control and keep spreading | 1 (2.9%) | 20 (6.4%) | 36 (10.5%) | 102 (32.2%) | 0.000 |
| High level of fear of family being infected | 12 (34.3%) | 162 (52.1%) | 222 (64.7%) | 253 (79.8%) | 0.000 |
| Felt worried about other health problems | 4 (11.4%) | 25 (8.0%) | 59 (17.2%) | 110 (34.7%) | 0.000 |
| Felt worried about family's other health problems | 10 (28.6%) | 115 (37.0%) | 169 (49.3%) | 224 (70.7%) | 0.000 |
| Felt worried about their or their family's finances | 11 (31.4%) | 130 (41.8%) | 198 (57.7%) | 230 (72.6%) | 0.000 |
| High level of fear of being quarantined | 5 (14.3%) | 68 (21.9%) | 112 (32.7%) | 160 (50.5%) | 0.000 |
| Felt worried about family/friends distancing themselves from me due to my job | 2 (5.7%) | 47 (15.1%) | 86 (25.1%) | 158 (49.8%) | 0.000 |
| High level of fear of being assigned to a Covid-19 ward | 1 (2.9%) | 42 (13.5%) | 84 (24.5%) | 131 (41.3%) | 0.000 |
| Felt reluctant to go to work | 2 (5.7%) | 9 (2.9%) | 40 (11.7%) | 118 (37.2%) | 0.000 |
| Felt worried about workload increasing | 1 (2.9%) | 40 (12.9%) | 123 (35.9%) | 197 (62.2%) | 0.000 |

In multivariable ordinal logistic regression results (Table 3), older age and being male continued to be significantly associated with lower distress levels, as were the following factors: feeling that the institution had effective protective measures in place, and reporting being satisfied at work. Conversely, suffering from atleast one symptom of burnout; reporting functional problems due to sleep-related issues (in the past 7 days); reporting high level of exhaustion (in the past 7 days); being a pharmacist (relative to a physician) and working in a tertiary cancer center; harboring fear about the virus spreading uncontrollably; fear that the virus threatened life; fear of alienation from family and friends; and fear of workload increases, were all significantly associated with reporting higher distress levels.

**Table 3.** Multivariable ordinal logistic regression examining the association between demographic, psychological and professional characteristics on distress level in a sample of Jordanian healthcare practitioners*.

|  | **Odds Ratio** | **p-value** | **95% confidence interval** |  |
| --- | --- | --- | --- | --- |
| **Age (years)\*\*** | 0.96 | 0.000 | 0.94 | 0.98 |
| **Being male (reference: female)\*\*** | 0.53 | 0.000 | 0.39 | 0.73 |
| **Live with spouse (reference: those living without a partner)** | 0.96 | 0.860 | 0.57 | 1.59 |
| **Live with older - 65 or older - person (reference: those not living with an older person)** | 1.00 | 0.986 | 0.75 | 1.34 |
| **Live with younger - 18 or younger - person (reference: those not living with younger people)** | 1.09 | 0.686 | 0.71 | 1.69 |
| **Have a child (reference: those without a child)** | 1.00 | 0.987 | 0.57 | 1.74 |
| **Educational level – Bachelors (reference)** |  |  |  |  |
| Educational level – Diploma | 0.83 | 0.400 | 0.54 | 1.28 |
| Educational level – Masters | 1.18 | 0.471 | 0.75 | 1.87 |
| **Profession – Physician (reference)** |  |  |  |  |
| Profession – pharmacist** | 2.36 | 0.029 | 1.09 | 5.10 |
| Profession – nurse | 1.00 | 0.994 | 0.62 | 1.61 |
| **Type of institution – non-cancer/general hospital (reference)** |  |  |  |  |
| Tertiary cancer center** | 1.63 | 0.004 | 1.17 | 2.29 |
| Community pharmacy | 0.67 | 0.309 | 0.31 | 1.46 |
| **Worked in a Covid ward (reference: those who worked in all other wards or did not work in hospital)** | 1.09 | 0.687 | 0.71 | 1.69 |
| **Worked with potential or suspected Covid-19 patients (reference: those who did not report working with suspected patients)** | 0.94 | 0.702 | 0.68 | 1.30 |
| **Experienced a high workload during outbreak (reference: those reporting moderate or low workload)** | 0.96 | 0.795 | 0.68 | 1.34 |
| **Reported atleast one symptom of burnout (reference: reported no symptoms of burnout)\*\*** | 3.02 | 0.000 | 2.10 | 4.35 |
| **Had [quite a bit, very much] exhaustion (reference: those who reported some or none)\*\*** | 2.46 | 0.000 | 1.72 | 3.52 |
| **Experienced [quite a bit, very much] sleep disturbances (reference: those who reported some or none)\*\*** | 2.35 | 0.000 | 1.65 | 3.34 |
| **Agreed that they were satisfied with work (reference: those who disagreed or were neutral to the statement)\*\*** | 0.33 | 0.000 | 0.23 | 0.47 |
| **Fear of infecting others (reference: those who reported no fear or little/some fear only)** | 1.13 | 0.529 | 0.77 | 1.68 |
| **Fear that life was under threat (reference: those who reported no fear or little/some fear only)\*\*** | 1.60 | 0.013 | 1.10 | 2.31 |
| **Fear that virus going out of control and continuing to spread (reference: those who reported no fear or little/some fear only)\*\*** | 2.03 | 0.003 | 1.26 | 3.26 |
| **Fear of workload increasing (reference: those who reported no fear or little/some fear only)\*\*** | 1.55 | 0.019 | 1.07 | 2.24 |
| **Fear of family/friends distancing themselves from respondent (reference: those who reported no fear or little/some fear only)\*\*** | 1.47 | 0.039 | 1.02 | 2.12 |
| **Fear of being assigned to a Covid-19 ward (reference: those who reported no fear or little/some fear only)** | 1.21 | 0.325 | 0.83 | 1.76 |
| **Agreed that N95 masks were available (reference: those who disagreed or were neutral to the statement)** | 1.14 | 0.438 | 0.82 | 1.57 |
| **Agreed that surgical masks were available (reference: those who disagreed or were neutral to the statement)** | 1.03 | 0.873 | 0.69 | 1.54 |
| **Agreed that protective measures were in place in institution (reference: those who disagreed or were neutral to the statement)\*\*** | 0.68 | 0.029 | 0.48 | 0.96 |
| **Reported confidence in the protective effect of surgical masks (reference: those who reported moderate to low confidence)** | 0.95 | 0.742 | 0.69 | 1.31 |
| **Reported confidence in the protective effect of N95 masks (reference: those who reported moderate to low confidence)** | 0.74 | 0.067 | 0.53 | 1.02 |

*\*Nonmedical professions (n=69) were excluded from the analysis*
*\*\*p-value $\leq 0.05$*

### Other results: personal protective equipment across professions

Nurses reported the highest rates of PPE availability and confidence in putting on and taking off PPE. Nurses also were more likely to have received training for PPE use, although it is notable that receipt of PPE training was 50.5% across all professions (Table 4).

**Table 4.**
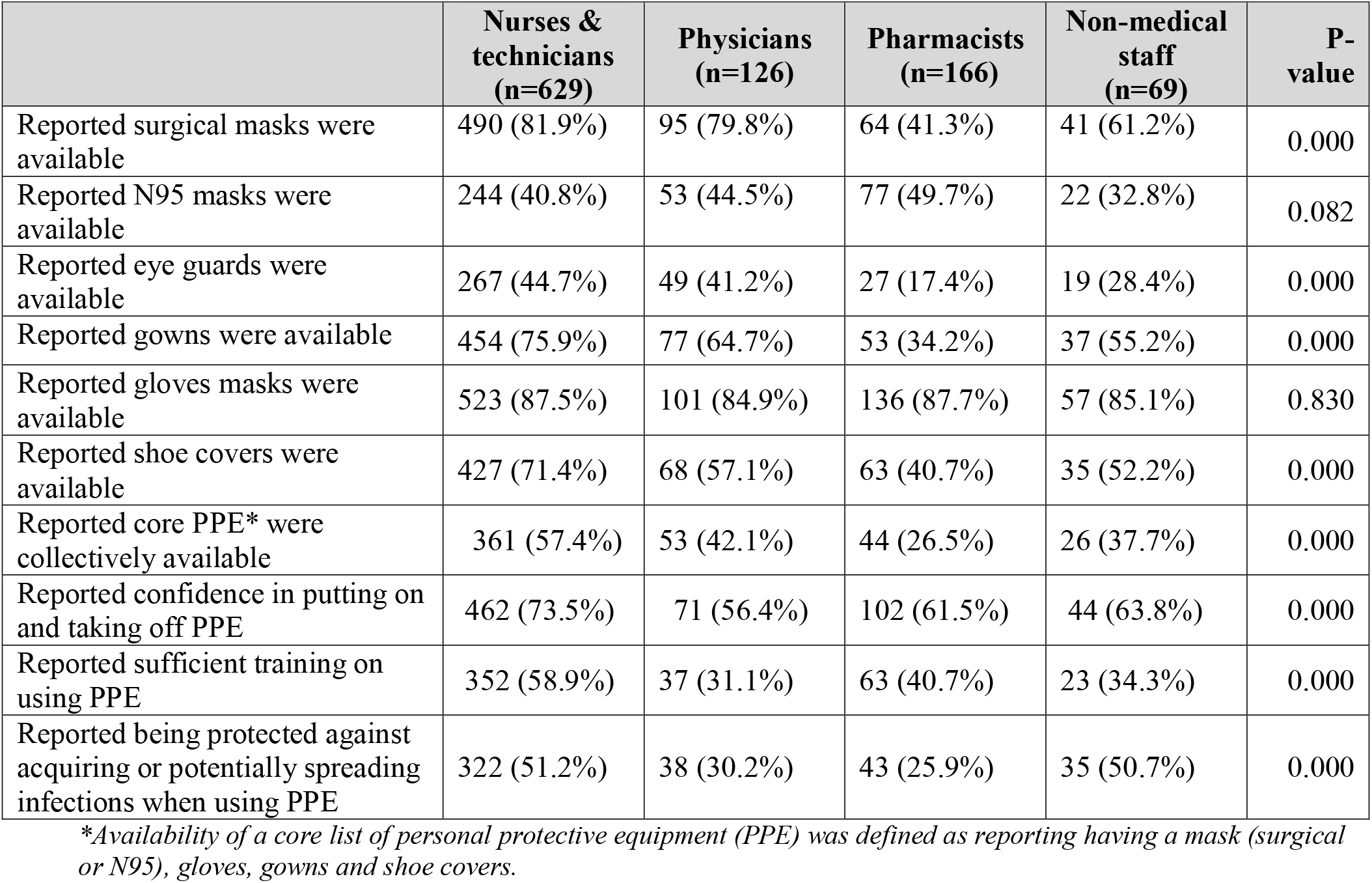
Access to personal protective equipment (PPE) and other perceptions related to PPE use across different professions (n=1,006). Column total percentages presented.

## DISCUSSION

Our study evaluates distress levels among healthcare providers in a country with a low caseload and under lockdown. Our data confirm significant fears and distress among healthcare employees – physicians, nurses, pharmacists, technicians and even nonmedical staff – working in healthcare facilities in Jordan during the lockdown. Thus, even in circumstances where caseloads were low, and the healthcare sector did not suffer from a severe stretching of resources (staff and PPE), distress and anxiety levels were considerable. Close to 32% of our sample reported high distress levels during the study period, with roughly 20% falling in the severe distress category. Approximately a third and a quarter also reported at least moderate anxiety and depression (respectively), and almost a third reported sleep problems and problems in functionality due to sleep issues. These numbers are comparable to other countries,(8, 10, 12, 36) and confirm the importance of tracking mental health in healthcare practitioners, even during periods of relative calm when caseloads are low. Our results suggest that we need to do more with regards to preparing and protecting our healthcare practitioners in anticipation of the realistic possibility of a future surge in Covid-19, given that our finding suggest that our practitioners are already predisposed and have experienced considerable psychological stress.

Our findings are valuable because they highlight specific demographic and professional factors that were more pronounced in those reporting high levels of distress. Older age (which we also considered a proxy for years of experience) was inversely associated with distress. Conversely, being female and working in a cancer hospital were significantly associated with a greater odds of being in a higher distress category. Others have reported similar findings with regards to gender.(8) With regards to working in a cancer setting, cancer centers are usually associated with high levels of burnout and distress.(37) Likely further aggravating this situation was the heightened concern regarding the potentially poor prognosis for cancer patients should they acquire Covid-19, and which has now been documented in other studies.(38) We also noted the emergence of hospital oncology pharmacists as a relatively distressed healthcare profession (levels of distress among them exceeded those found among other professions and their community counterparts). We had originally hypothesized that all pharmacists would experience greater distress, because during the lockdown period, pharmacies continued their operations, and community pharmacies in particular became the only accessible source of some basic healthcare services for the public. This was not observed. The heightened distress of hospital oncology pharmacists in particular may be explained by the fact that despite fewer pharmacists per shift, they continued to deliver outpatient medications to a greater number of vulnerable cancer patients (using delivery services, which in itself may have posed additional stress given a greater number of patients now not being counseled in the normal manner).

The study also highlighted several psychological and functional factors that were indicative of being in higher distress: reporting burnout, physical exhaustion, problems in functionality due to sleep issues; and harboring fears specifically pertaining to the virus spreading beyond control, being alienated from friends or family, and increasing workloads. On the other hand, general satisfaction at work correlated with lower distress levels. The psychological and functional factors that emerged in our analysis are useful in highlighting thoughts as well as concerns that, if expressed by employees in their clinical practice, can prompt leaders in the workplace to take notice and explore the possibility of distress as well as attempt to alleviate it early on. For example, continually tracking burnout as well as functionality due to sleep issues or exhaustion may be of potential use in preempting healthcare workers’ reaching high levels of distress.

Fostering a working environment that ensures worker satisfaction also may be an additional means of buffering against distress.

In our sample, the most widely resonant fear was fear of infecting a family member or a colleague (83% were concerned about this, whereas only 33% indicated they were concerned about being infected). This is similar to what others have noted:(9, 39) Fear of infecting others was likely more prominent in our sample due to the cultural setting: in Jordan, similar to other countries in the Middle East, long-term care facilities such as nursing homes, skilled nursing facilities, and assisted living facilities, are scarce. Elderly people are usually cared for by their families who typically live with them or very close by. Furthermore, it is unusual for young unmarried adults to live alone. Thus, it is common to find Jordanian households with both young and old family members (and relatively large family units), likely explaining why the majority of healthcare professionals were concerned about infecting others.

With regards to PPE, although on a bivariate level, their availability was associated with distress, it is noteworthy that availability of PPE did not exert a significant effect on distress in our multivariable model. This direction is contrary to what has been seen in high caseload countries, where PPE and ventilator shortages have been faced,(32) and where difficult and stressful decisions have had to be made that further increased healthcare practitioners’ distress. Our finding is not surprising, given that the country did not suffer from a surge in cases, and even within the cases reported, the majority were mild to moderate cases that did not require critical care. Nevertheless, the proportions of respondents who stated they were trained in PPE use was relatively low (especially among physicians), which raises concerns about preparedness of healthcare workers should a future outbreak occur.

Our study has some limitations. We were not able to qualitatively examine in an in-depth manner the exact sources of distress among our high-distress sample, and how these interacted with one another within individuals (others in high caseload settings have used interviews in limited samples to detail specific sources of distress(39)). We also speculate that a source of distress for healthcare workers that was not probed in our study was the general experience of the stringent lockdown. Our survey was not designed to specifically measure this, but others have shown that generally experiencing a lockdown and quarantines negatively impacts mental health.(40) Furthermore, our survey was cross-sectional in nature, and did not capture the effect of fluctuations in the general Covid-19 situation on distress. However, it is relevant to note that although there is a possibility that the symptoms we report may have existed prior to the Covid-19 situation, we additionally inquired about whether or not – among those who reported any symptoms of anxiety or depression, and those reporting sleep issues or exhaustion – such symptoms existed pre-Covid-19, and found that less than 17% of respondents reported that they suffered from these symptoms in the same intensity pre-Covid-19.

Despite its limitations, we have been able to collect valuable data on a large and diverse sample of medical professionals representing various healthcare facilities (governmental and academic hospitals including a tertiary cancer center, and community-based pharmacies), and within a critical time period, during and shortly after a lockdown. Our results will help in identifying which potential healthcare professionals to target as well as specific topics to discuss, in order to preempt workers reaching a state of high distress in the medical workplace, thus preparing them to handle the Covid-19 situation with resilience, regardless of the continually changing environment and the potential for caseload changes in the future. Follow-up studies will be of value in tracking medical practitioners’ responses in the face of the unpredictable Covid-19 situation.

## Data Availability

Data are available upon reasonable request from the Principal Investigator.

## References

1. Ho SM, Kwong-Lo RS, Mak CW, Wong JS. Fear of severe acute respiratory syndrome (SARS) among health care workers. Journal of consulting and clinical psychology. 2005;73(2):344–9.

2. Wong TW, Yau JK, Chan CL, Kwong RS, Ho SM, Lau CC, et al. The psychological impact of severe acute respiratory syndrome outbreak on healthcare workers in emergency departments and how they cope. European journal of emergency medicine : official journal of the European Society for Emergency Medicine. 2005;12(1):13–8.

3. Lancee WJ, Maunder RG, Goldbloom DS. Prevalence of psychiatric disorders among Toronto hospital workers one to two years after the SARS outbreak. Psychiatric services (Washington, DC). 2008;59(1):91–5.

4. Koh D, Lim MK, Chia SE, Ko SM, Qian F, Ng V, et al. Risk perception and impact of Severe Acute Respiratory Syndrome (SARS) on work and personal lives of healthcare workers in Singapore: what can we learn? Medical care. 2005;43(7):676–82.

5. Goulia P, Mantas C, Dimitroula D, Mantis D, Hyphantis T. General hospital staff worries, perceived sufficiency of information and associated psychological distress during the A/H1N1 influenza pandemic. BMC infectious diseases. 2010;10:322.

6. McAlonan GM, Lee AM, Cheung V, Cheung C, Tsang KW, Sham PC, et al. Immediate and sustained psychological impact of an emerging infectious disease outbreak on health care workers. Canadian journal of psychiatry Revue canadienne de psychiatrie. 2007;52(4):241–7.

7. Thombs BD, Bonardi O, Rice DB, Boruff JT, Azar M, He C, et al. Curating evidence on mental health during COVID-19: A living systematic review. J Psychosom Res. 2020:110113.

8. Pappa S, Ntella V, Giannakas T, Giannakoulis VG, Papoutsi E, Katsaounou P. revalence of depression, anxiety, and insomnia among healthcare workers during the COVID-19 pandemic: A systematic review and meta-analysis. Brain Behav Immun. 2020.

9. SCCM COVID-19 Rapid-Cycle Survey 2 Report. https://sccm.org/Blog/May-2020/SCCM-COVID-19-Rapid-Cycle-Survey-2-Report?_zs=NIUjd1&_zl=xI8l6.

10. Zhang SX, Liu J, Afshar Jahanshahi A, Nawaser K, Yousefi A, Li J, et al. At the height of the storm: Healthcare staff’s health conditions and job satisfaction and their associated predictors during the epidemic peak of COVID-19. Brain Behav Immun. 2020.

11. Simione L, Gnagnarella C. PREPRINT. Differences between health workers and general population in risk perception, behaviors, and psychological distress related to COVID-19 spread in Italy. 2020.

12. Rossi R, Socci V, Pacitti F, Di Lorenzo G, Di Marco A, Siracusano A, et al. Mental Health Outcomes Among Frontline and Second-Line Health Care Workers During the Coronavirus Disease 2019 (COVID-19) Pandemic in Italy. JAMA network open. 2020;3(5):e2010185.

13. Alqutob R, Al Nsour M, Tarawneh MR, Ajlouni M, Khader Y, Aqel I, et al. COVID-19 crisis in Jordan: Response, scenarios, strategies, and recommendations. JMIR Public Health Surveill. 2020.

14. https://corona.moh.gov.jo/ar.

15. Basheti IA, Nassar R, Barakat M, Alqudah R, Abufarha R, Mukattash TL, et al. Pharmacists’ readiness to deal with the coronavirus pandemic: Assessing awareness and perception of roles. Res Social Adm Pharm. 2020.

16. Khader Y, Al Nsour M, Al-Batayneh OB, Saadeh R, Bashier H, Alfaqih M, et al. Dentists’ Awareness, Perception, and Attitude Regarding COVID-19 and Infection Control: Cross-Sectional Study Among Jordanian Dentists. JMIR Public Health Surveill. 2020;6(2):e18798.

17. Suleiman A, Bsisu I, Guzu H, Santarisi A, Alsatari M, Abbad A, et al. Preparedness of Frontline Doctors in Jordan Healthcare Facilities to COVID-19 Outbreak. Int J Environ Res Public Health. 2020;17(9).

18. Naser AY, Dahmash EZ, Al-Rousan R, Alwafi H, Alrawashdeh HM, Ghoul I, et al. PREPRINT. Mental health status of the general population, healthcare professionals, and university students during 2019 coronavirus disease outbreak in Jordan: a cross-sectional study. 2020.

19. Greenberg N, Docherty M, Gnanapragasam S, Wessely S. Managing mental health challenges faced by healthcare workers during covid-19 pandemic. BMJ (Clinical research ed). 2020;368:m1211.

20. Walton M, Murray E, Christian MD. Mental health care for medical staff and affiliated healthcare workers during the COVID-19 pandemic. Eur Heart J Acute Cardiovasc Care. 2020:2048872620922795.

21. Kessler RC, Andrews G, Colpe LJ, Hiripi E, Mroczek DK, Normand SL, et al. Short screening scales to monitor population prevalences and trends in non-specific psychological distress. Psychological medicine. 2002;32(6):959–76.

22. Dolan ED, Mohr D, Lempa M, Joos S, Fihn SD, Nelson KM, et al. Using a single item to measure burnout in primary care staff: a psychometric evaluation. Journal of general internal medicine. 2015;30(5):582–7.

23. Patient-Reported Outcomes Measurement Information System (PROMIS) Health Measures: Anxiety Short Form 4a. Available from http://www.healthmeasures.net/explore-measurement-systems/promis.

24. Patient-Reported Outcomes Measurement Information System (PROMIS) Health Measures: Emotional Distress-Depression – Short Form 4a. Available from http://www.healthmeasures.net/explore-measurement-systems/promis.

25. Patient-Reported Outcomes Measurement Information System (PROMIS) Health Measures: Sleep impact short-form. Available from http://www.healthmeasures.net/explore-measurement-systems/promis.

26. Patient-Reported Outcomes Measurement Information System (PROMIS) Health Measures. Sleep-related impairment (short-form). Available from http://www.healthmeasures.net/explore-measurement-systems/promis.

27. Patient-Reported Outcomes Measurement Information System (PROMIS) Health Measures. Fatigue (short-form). Available from http://www.healthmeasures.net/explore-measurement-systems/promis.

28. Forman-Hoffman VL, Muhuri PK, Novak SP, Pemberton MR, Ault KL, Mannix D. Center for Behavioral Health Statistics and Quality, SAMHSA: CBHSQ Data Review, August 2014. Psychological Distress and Mortality among Adults in the U.S. Household Population. https://www.samhsa.gov/data/sites/default/files/CBHSQ-DR-C11-MI-Mortality-2014/CBHSQ-DR-C11-MI-Mortality-2014.pdf. 2014.

29. Schalet BD, Cook KF, Choi SW, Cella D. Establishing a common metric for self-reported anxiety: linking the MASQ, PANAS, and GAD-7 to PROMIS Anxiety. J Anxiety Disord. 2014;28(1):88–96.

30. Choi SW, Schalet B, Cook KF, Cella D. Establishing a common metric for depressive symptoms: linking the BDI-II, CES-D, and PHQ-9 to PROMIS depression. Psychol Assess. 2014;26(2):513–27.

31. Friedberg, M. W., Chen, P. G., Van Busum K. R., Aunon, F., Pham, C., Caloyeras, J. P., Mattke, S., Pitchforth, E., Quigley, D. D., Brook, R. H., Crosson, F. H., Tutty, M. Factors Affecting Physician Professional Satisfaction and Their Implications for Patient Care, Health Systems, and Health Policy. Santa Monica, CA: RAND Corporation, 2013. https://www.rand.org/pubs/research_reports/RR439.html. Also available in print form.

32. World Health Organization, March 3, 2020 News Release. Shortage of personal protective equipment endangering health workers worldwide. https://www.who.int/news-room/detail/03-03-2020-shortage-of-personal-protective-equipment-endangering-health-workers-worldwide.

33. Imai T, Takahashi K, Hasegawa N, Lim MK, Koh D. SARS risk perceptions in healthcare workers, Japan. xmerging infectious diseases. 2005;11(3):404–10.

34. Shah, A. (2018). ASDOC: Stata module to create high-quality tables in MS Word from Stata output. Statistical Software Components S458466, Boston College Department of Economics.

35. StataCorp. 2019. Stata Statistical Software: Release 16. College Station, TX: StataCorp LLC.

36. Tan BYQ, Chew NWS, Lee GKH, Jing M, Goh Y, Yeo LLL, et al. Psychological Impact of the COVID-19 Pandemic on Health Care Workers in Singapore.Annals of internal medicine. 2020.

37. Hlubocky FJ, Back AL, Shanafelt TD. Addressing Burnout in Oncology: Why Cancer Care Clinicians Are At Risk, What Individuals Can Do, and How Organizations Can Respond. Am Soc Clin Oncol Educ Book. 2016;35:271–9.

38. Kuderer NM, Choueiri TK, Shah DP, Shyr Y, Rubinstein SM, Rivera DR, et al. Clinical impact of COVID-19 on patients with cancer (CCC19): a cohort study. Lancet (London, England). 2020.

39. Chen Q, Liang M, Li Y, Guo J, Fei D, Wang L, et al. Mental health care for medical staff in China during the COVID-19 outbreak. The lancet Psychiatry. 2020;7(4):e15–e6.

40. Brooks SK, Webster RK, Smith LE, Woodland L, Wessely S, Greenberg N, et al. The psychological impact of quarantine and how to reduce it: rapid review of the evidence. The Lancet. 2020;395(10227):912–20.

